# Effect of *Bacillus coagulans* Unique IS-2 in Inflammatory Bowel Disease (IBD): A Randomized Controlled Trial

**DOI:** 10.1101/2021.07.18.21260556

**Authors:** V Deepak Bamola, Divya Dubey, Projoyita Samanta, Saurabh Kedia, Ratna Sudha Madempudi, Jayanthi Neelamraju, Vineet Ahuja, Rama Chaudhry

## Abstract

Probiotic intervention is an important approach for the treatment and health restoration in inflammatory bowel disease (IBD). A study was conducted to assess the effect of *Bacillus coagulans* Unique IS-2 in IBD patients. Recruited subjects were given either probiotic *B. coagulans* Unique IS-2 or placebo for 4 weeks as per randomization. Survival of the given probiotic strain in GI, presence of beneficial gut bacteria, serum cytokines, serum serotonin and serum dopamine, symptoms of disease, physical, behavioral and psychological parameters of the subjects were evaluated before and after intervention. In this study *B. coagulans* Unique IS-2 was well tolerated with no severe adverse events in IBD patients and demonstrated good survival in GI tract by significantly high detection in probiotic treated group (p <0.001). Significant enhancement in beneficial *Lactobacilli* was observed in probiotic treated group (p <0.01). Metagenomic analysis also showed an increase in the abundance of bacterial genera *Bacillus, Lactobacillus, Bifidobacterium, Faecalibacterium, Bacteroides, Megamonas, Lachnospira, Blautia*, Alistipes and decrease in *Sutterella, Dialister, Roseburia and Megasphaera* was observed in the post intervention samples in the treatment group. Increased secretion of cytokine IL-10 and variable decrease in the secretion of IL-6, IL-1β, TNF-α, IL -17 and IL -23 was observed in in the probiotic treated group. Post intervention change in serum serotonin and serum dopamine was not significant in both the groups. A reduction in the severity of disease symptoms and improvement in the physical, behavioral and psychological parameter was observed in the probiotic treated group. The observed results demonstrated that *B. coagulans* Unique IS-2 with SMT was effective in adult IBD patients. Study was registered with Clinical Trials Registry India (CTRI) - (*registration no*.*-CTRI/2019/11/022087*).

## Introduction

Inflammatory bowel disease (IBD) is a gastrointestinal (GI) disease characterized by chronic inflammation. The incidence of IBD is rising all over the world^1^ including Asia^2^. Across the globe India is projected to have one of the highest IBD burden in spite of having lower prevalence as compared to the West^3,4^. IBD significantly diminishes the quality of life of the affected individuals by disturbing health, daily work, education, and social relationships which cause a considerable impact on health and economy^5^. Etiology and pathogenesis of IBD is multi-factorial and involves genetics, ethnicity, diet, lifestyle, environmental factors, immunity and gut microbiota of the individual^6,7^. Higher stress level and disturbance in psychology in IBD patients may trigger the inflammation and increases the severity of disease^8^.

The human gut microbiota has demonstrated physiological functions associated with homeostasis, nutrition, immunity and defense of the host. Normally the gut maintains a homeostatic condition but in IBD this homeostasis gets disturbed and leads to intestinal inflammation^9^. Down-regulation of the immune responses may allow the damaged site to heal and reset the normal physiological functions^7^. Alteration in the gut microbiota, referred to as dysbiosis, plays a key role in the pathogenesis of IBD^10,11^. Dysbiosis may occur in both inflamed and non-inflamed areas in patients^12^ and is well recognized sign of IBD^13^. Modification of gut microbiota with probiotic intervention to attenuate inflammatory activity and prevent relapses in IBD are considered as an important approach for treatment and health restoration in IBD^11,14^ by relieving intestinal dysbiosis and GI inflammation^10^.

GI disorders modulate the gut function and influence the emotional and cognitive factors including mood, anxiety, pain and negative effects, decision making, restlessness. This change in emotional and cognitive factors may be associated with serum serotonin and dopamine in IBD patients^15^. The lowering of serotonin levels can result in mood disorders, such as anxiety or depression and hence consider playing an important role in regulating physical and psychological symptoms^16^ and activation of immune response and gut inflammation^15^. Dopamine is an important neurotransmitter which affects behavior, psychology, immune functions and GI functions and its unregulated production in IBD is already reported^17^. Evaluation of these factors has turn out to be an important factor to monitor the severity of the disease. Gut microbiota and probiotics have demonstrated the effect on behavior, psychology, mood, and cognition^18,19^.

*Bacillus* strains are stable at room temperature and hence gaining a lot of attention. *B. coagulans* is a spore-forming, gram-positive, non-pathogenic, facultative anaerobic, lactic acid-producing bacteria and resistant to high temperatures^20^. *B. coagulans* Qualified Presumption of Safety (QPS) list^21^ and has been reported Generally Recognized as Safe (GRAS) and considered safe by European Union Food Safety Authority (EFSA) and US Food and Drug Administration (FDA). *B coagulans* Unique IS-2 (ATCC PTA-11748, MTCC 5260) is a shelf-stable, resistant to bile acids and acidic conditions of the stomach, clinically established probiotic strain and with proven safety and efficacy in the treatment of constipation, diarrhea^22–25^ bacterial vaginosis^26^ and irritable bowel syndrome (IBS)^27^. In this study we assessed the effect of *B coagulans* Unique IS-2 on adults with inflammatory bowel disease (IBD) receiving standard medical treatment (SMT). Survival of the given probiotic *B coagulans* Unique IS2 in the gut, presence of beneficial gut bacteria, serum cytokines, serotonin and dopamine, IBD symptoms, physical and psychological parameters were evaluated before and after intervention.

## Methodology

### Study design

Randomized, double blind, placebo controlled trial. Study was registered with Clinical Trials Registry (CTRI) India (CTRI/2019/11/022087).

### Site

Outpatients from a tertiary care hospital.

### Ethical approval and written informed consent

Ethical approval for the study was obtained from Institute Ethics Committee of AIIMS, New Delhi, India (Ref – IEC.478/07.10.2016.OP-7). Expected duration of participation, expected benefits, associated risks, and maintenance of confidentiality of records were explained to each participant and a written informed consent was obtained before enrollment in the study.

### Subject / Selection of patients

Clinically diagnosed adult patients of ulcerative colitis (UC) with mild to moderate severity between age group (18–60 years) under standard Medical treatment (SMT) patients were included. Simple Clinical Colitis Activity Index (SCCAI) score^28^ was used to quantify UC disease activity. SCCAI score was calculated by evaluating different disease symptoms including bowel frequency, urgency of defecation, blood in stool, abdominal cramps and general wellbeing of the patients. SMT for the enrolled patient in this study was 5-aminosalicylic acid (5-ASA) - Sulfasalazine (3 grams/ day) or Mesalamine 800 mg orally 3 times a day.

### Inclusion criteria

(a) Adult patient clinically diagnosed with Ulcerative Colitis (UC), (b) patients of either sex of the age range from 18-60 year, (d) patient ready to participate in the study and giving written consent, (e) patient visiting to Out Patient Department (OPD) of AIIMS for treatment.

### Exclusion criteria

(a) Patient diagnosed with any kind of carcinoma, (b) Patient diagnosed with any other gastrointestinal disease, (c) Patient suffering from Immunodeficiency disorder, (d) Patient is taking any probiotic drug/ or having consumed probiotic in the last one month, (e) patient not taking food through oral route, (f) Patient having undergone any kind of gastrointestinal surgery in the last three month.

### Enrolment of patients

After establishing the eligibility on screening, a total of 100 patients were recruited and randomized. Patients were called for baseline visit (day 0). Medical history, medications, physical examination and vital signs were assessed during hospital visit. *B. coagulans* Unique IS-2 (2billion-CFU/capsule) twice in day (total 4 billion CFU / per day) or placebo (matching in size and appearance, contained only excipient, maltodextrin) twice in a day was given to qualified patients for 4 weeks followed by observation and telephonic follow-up of dose compliance.

### Sample size determination

Statistical software STATA (Version 14, USA) was used for sample calculation. To identify presence of proportion difference the assumption was made that minimum of 118 subjects required to be screened and 94 patients required assessing the endpoint in the study which will reject the null hypothesis.

### Intervention

Fully characterized FDA / DCGI/ FSSAI approved probiotic strain *Bacillus coagulans* Unique IS-2 (ATCC PTA-11748, MTCC 5260) was used as an intervention agent in this study for 4 weeks along with placebo as per randomization. The patients were explained to consume one capsule post meal twice a day for 4 weeks along with the SMT. The compliance was ensured by telephonic follow-up and scheduled hospital visits.

### Randomization

Computerized permuted blocks randomization was done in 1:1 ratio and generated by nQuery clinical trial design platform / Sample Size Software. It consisted of two phases: screening, baseline visit 1 (week 0 / day 0), visit 2 (1 week after completing intervention / week 5). The randomization codes were kept blinded.

### Outcome measures

The efficacy outcomes were measured after administration by (i) detection of *Bacillus coagulans* Unique IS-2, (ii) Change in beneficial Lactobacillus and other gut microbiota, (iii) Change in pro and anti-inflammatory cytokines, (iv) Change in symptoms of the disease, (v) Change in serum concentration of serotonin and dopamine, (vi) Changes in physical and psychological parameters. Each participant was requested to answer the designed questionnaire for the assessment of physical, behavioral and psychological parameters as per the Hopkins Symptom Checklist (HSCL): A self-report symptom inventory^29,30^. Physical parameters including muscle stiffness, heartburn, headache, shakiness, sleep problem, difficulty in completing work, procrastination, overwhelming, feeling of depression, trouble relaxing, nervousness, poor concentration, restlessness and quick temper were evaluated. Parameters were evaluated based on scores, the decrease in the score indicating the reduction in the severity of symptoms and increase in the score indicating an augmentation in the severity of the symptom.

### Safety evaluation

Safety of investigational product was assessed by adverse event reporting. During hospital visit of the patient physical examination, monitoring of vital signs and routine laboratory investigations.

### Sample collection and processing

A stool sample in a sterile container and a blood sample in a plain vial were collected from each enrolled subject before and after intervention. Stool samples were aliquoted and processed for microbial identification and bacterial DNA isolation. Blood samples were processed for serum separation for cytokine assays, serotonin and dopamine concentration.

### Microbial culture and identification

*B coagulans* Unique IS-2 and *Lactobacillus spp*. were checked in each patient before and after intervention using different bacterial media. Mueller Hinton (MH) broth and agar (Difco Laboratory, Detroit, MI) was used for the cultivation of *Bacillus* strains. Sample were incubated for 24 hours at 37°C in MH broth and then plated on MH agar plate. de Man, Rogosa and Sharpe (MRS) broth and agar (Difco Laboratory, Detroit, MI) was used to grow *Lactobacillus*. The stool sample were incubated for 48 hours at 37°C in MRS broth in Anaerobic Glove Box (Anaerobic Workstation-Whitley DG250-DonWhitley Scientific, United Kingdom) in anaerobic condition and then plated on MRS agar plate. The isolated colonies were identified by standard culture and biochemical method, matrix-assisted laser desorption / ionization-(MALDI-and the mass analyzer is time-of-flight (TOF) analyzer (bioMérieuxInc, USA) and molecular method.

### Molecular identification of Bacteria

*B. coagulans* was identified via 16S rRNA sequencing using published primers: forward 5’-ACAGGGCTTTCAGATACCCG-3’ and reverse 5’-CGGGGATCCGTCCATCAAAA-3’. Sequence similarity was checked using BLAST, NCBI and it was 96% identical. A known strain of *B. coagulans* Unique IS-2 was used as a positive control. The reaction mixture consisted of 0.5µl of dNTP (10mM), 0.5ul of DNA template (177ng/µl), 2.5 µl of reaction buffer (10X) with MgCL2, 0.5ul of each of primers (pm/µl), 0.5µl of 5U/µlTaq DNA polymerase (Thermo Scientific, USA) and 20 µl of nuclease free H_2_O. Denaturation was done at 94°C for 5min, followed by 30 cycles consisting of, 94°C for 1 min, 56°C for 1 min, and 72°C for 1 min which has been ended by a final amplification step at 72°C for 8min, using the PCR machine (Applied Biosystems, USA). PCR product was analyzed by the electrophoresis in 1% agarose gel and gel bands were observed and recorded using via Gel Doc System (BioRad, USA).

## Next generation sequencing (NGS) of Gut microbiota

### Fecal samples collection and DNA isolation

Fecal samples were collected in sterile container and processed for DNA isolation. Total DNA was extracted by using QIAamp DNA Stool Mini Kit (Qiagen) with some modification to increase DNA yield^31^. The quality and quantity of DNA was checked by Nanodrop (TECAN Nano quant). 16S rRNA amplicon sequencing was performed using Illumina MiSeq^®^ sequencing system (Illumina, San Diego, CA, USA).

### Sequencing Methodology

Bacterial 16S rRNA hyper variable regions V3-V4 were amplified using V3-V4F (CCTACGGGNGGCWGCAG) and V3-V4R (GACTACHVGGGTATCTAATCC) primers. 25ng of DNA was used for PCR amplification using KAPA HiFi HotStart Ready Mix. The PCR was performed with standard protocol and the amplicons were purified using Ampure beads to remove unused primers. The amplicon product was amplified with Illumina primers to generate sequencing libraries. Qubit dsDNA assay kit was used for libraries preparation. Sequencing was done using Illumina Miseq with 2×300PE sequencing kit. The sequence data quality was checked using FastQC and MultiQC software.

### Data Analysis

The analysis was done as per standard methodology^32^. Only QC passed reads were transferred into mothur and the pairs were aligned. The ambiguous contigs were rejected and duplicates were merged. Chimeric sequences were identified by a known reference and UCHIME algorithm was used. Using Silva v.132 database final filtered contigs were classified into taxonomical outlines and clustered into Operational Taxonomic Unit (OTUs) and abundance was calculated. Alpha diversity was assessed for richness and relative abundance of bacteria. Chao1 and ACE indices were used for richness and Shannon, Simpson, and Fisher were used for both richness and relative abundance. Kruskal-Wallis rank sum test was carried out to identify statistically significant difference among OTUs abundance between groups.

### ELISA for Cytokines, serum serotonin and dopamine

Serum samples were tested and quantified for IL10, IL6, IL17, IL23, IL 1β, TNF, serum serotonin and dopamine as per standard protocol and manufacture’s instruction (Fine Test, Fine Biotech Co. Ltd). In brief, standards and test samples were added to 96 well plates and incubated at 37^0^C for 90 min. After incubation, wells were washed with wash buffer and secondary antibody were added and incubated at 37°C for 60 min. Further, wells were washed and HRP-Streptavidin was added and incubated at 37^0^C for 30 min. Multiple washing was done with the wash buffer to wash unbound conjugates. TMB substrates were used to visualize HRP enzymatic reaction. Absorbance at 450 nm has been measured using the microplate reader (Nanodrop, Nanoquant Infinite M 200 Pro (Texan, Austria GmbH) and the concentration were calculated.

### Data analysis

Here we have reported the analysis of data of the patients of ulcerative colitis (UC). Data from 48 subjects in treatment group and 49 subjects in placebo group were analysed. Statistical analysis was done by STATA statistical software (Version 14, USA). Statistical evaluation of parameters was assessed by Chi-square / two sample *t-*test and p value < 0.05 was considered as statistically significant.

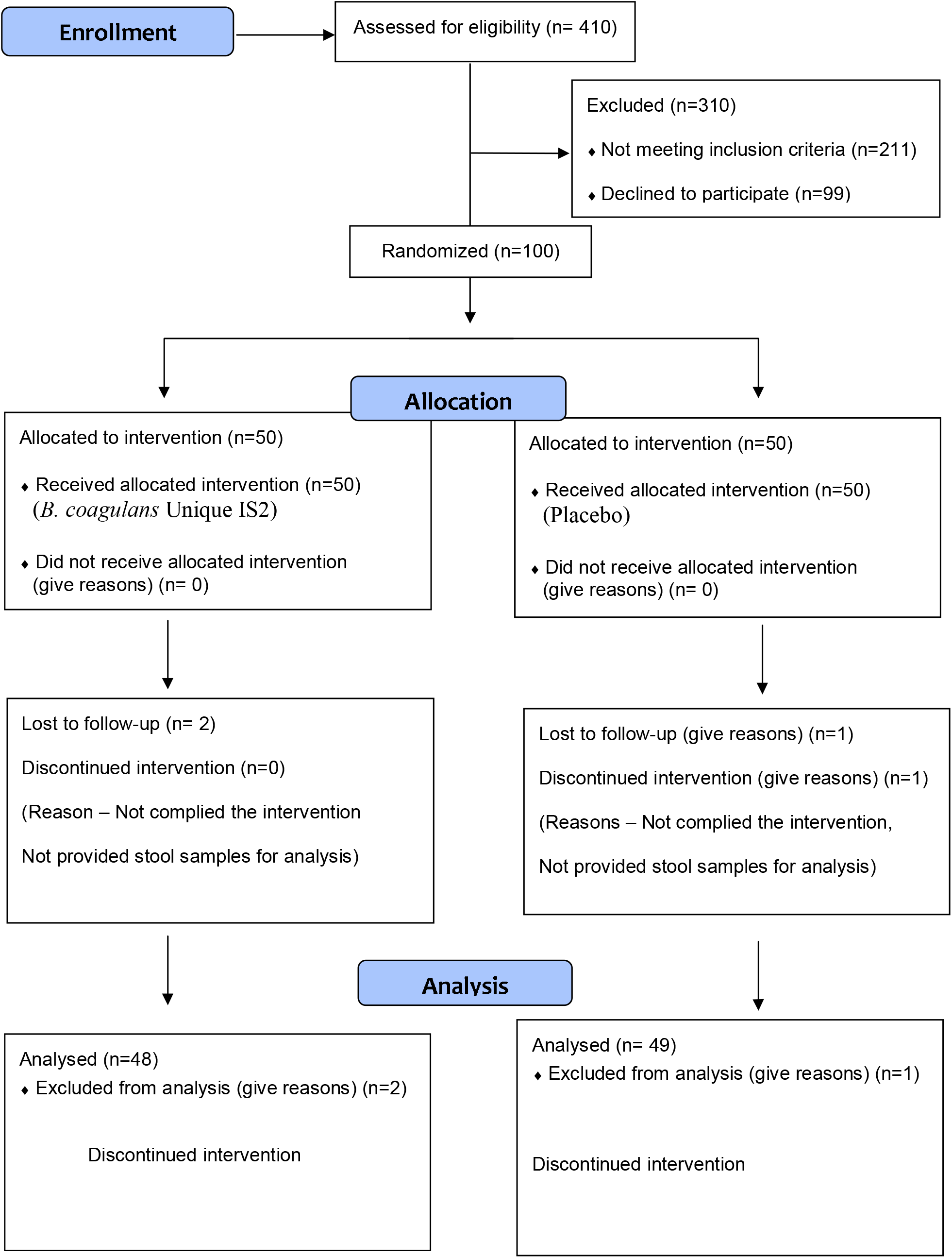
‘CONSORT’ Flow Chart’.

## Results

### Survival of *Bacillus coagulans* Unique IS2 in the gut

The samples of the enrolled subjects in the both treatment and placebo group were assessed for the presence of *Bacillus coagulans* Unique IS-2 before and after treatment. No *B coagulans* was detected in pre-treatment sample in both the groups. In the probiotic treated group *B coagulans* was detected in 65.21 % subjects by microbial culture method and 76.08 % subjects by molecular method which was significant as compared to placebo (p <0.0001). The samples from the subjects of placebo group were also assessed for the presence of *B coagulans* with no *B coagulans* being detected in placebo group by both microbial and molecular methods. These results indicate that the probiotic strain *B coagulans* Unique IS-2 was able to survive in GI tract of recruited IBD patients.

### Detection of beneficial Lactobacilli by microbial culture method

The samples of the enrolled IBD patients in both probiotic treated group and placebo group were assessed for the presence of beneficial Lactobacilli, before and after intervention. Results revealed that in the probiotic treated group, beneficial *Lactobacilli* were detected in 54.34 % and 78.26 % patients before and after intervention respectively. In the placebo group beneficial *Lactobacilli* were detected in 52.08 % and 47.91 % of patients before and after intervention respectively. The detection of beneficial *Lactobacilli* after intervention in the probiotic treated group was significantly high (p <0.01) which indicated that the probiotic strain *Bacillus coagulans* Unique IS-2 was able to enhance the presence of beneficial *Lactobacilli* in recruited IBD patients.

### Next generation Sequencing (NGS) and Metagenomic analysis of gut microbiota

#### Phyla

Operational Taxonomic Unit (OTUs) were calculated for different bacterial taxon including phyla, class, orders, families, genera and species before and after intervention in both probiotic treated and placebo group. Analysis revealed phylum Firmicutes Bacteriodetes, Proteobacteria, Actinobacteria, Euryarchaeota and Verrucomicrobia were abundant in both the groups. The average % OTUs of phylum *Firmicutes* were in 33.13% and 36.23% in pre and post intervention sample respectively in treatment group and 37.90% and 42.89% in placebo group. The average % OTUs of phylum *Bacteriodetes* were in 40.12% and 40.28% in pre and post intervention sample respectively in treatment group and 32.49% and 40.28% in pre and post intervention sample respectively in placebo group. OTUs of phylum *Bacteriodetes* were significantly increased in post intervention sample in placebo group. The average % OTUs of phylum *Proteobacteria* were in 12.90% and 7.50% in pre and post intervention sample respectively in treatment group and 16.75% and 6.21% in placebo group. The average % OTUs of phylum *Actinobacteria* were in 6.64% and 4.29% in pre and post intervention sample respectively in treatment group and 8.62% and 2.04% in placebo group. Decrease in OTUs of phylum *Proteobacteria* and phylum *Actinobacteria* was significant in the treatment group. The average % OTUs of phylum *Euryarchaeota* were in 1.02% and 2.36% in pre and post intervention sample respectively in treatment group and 2.03% and 0.55% in placebo group. OTUs of phylum Euryarchaeota were increased in post intervention sample in treatment group and decreased in placebo group. The average % OTUs of phylum Verrucomicrobia were in 0.01% and 0.04% in pre and post intervention sample respectively in treatment group and 0.96% and 0.10% in placebo group. OTUs of phylum Verrucomicrobia were increased in post intervention sample in treatment group and decreased in placebo group.

**Figure 1:**
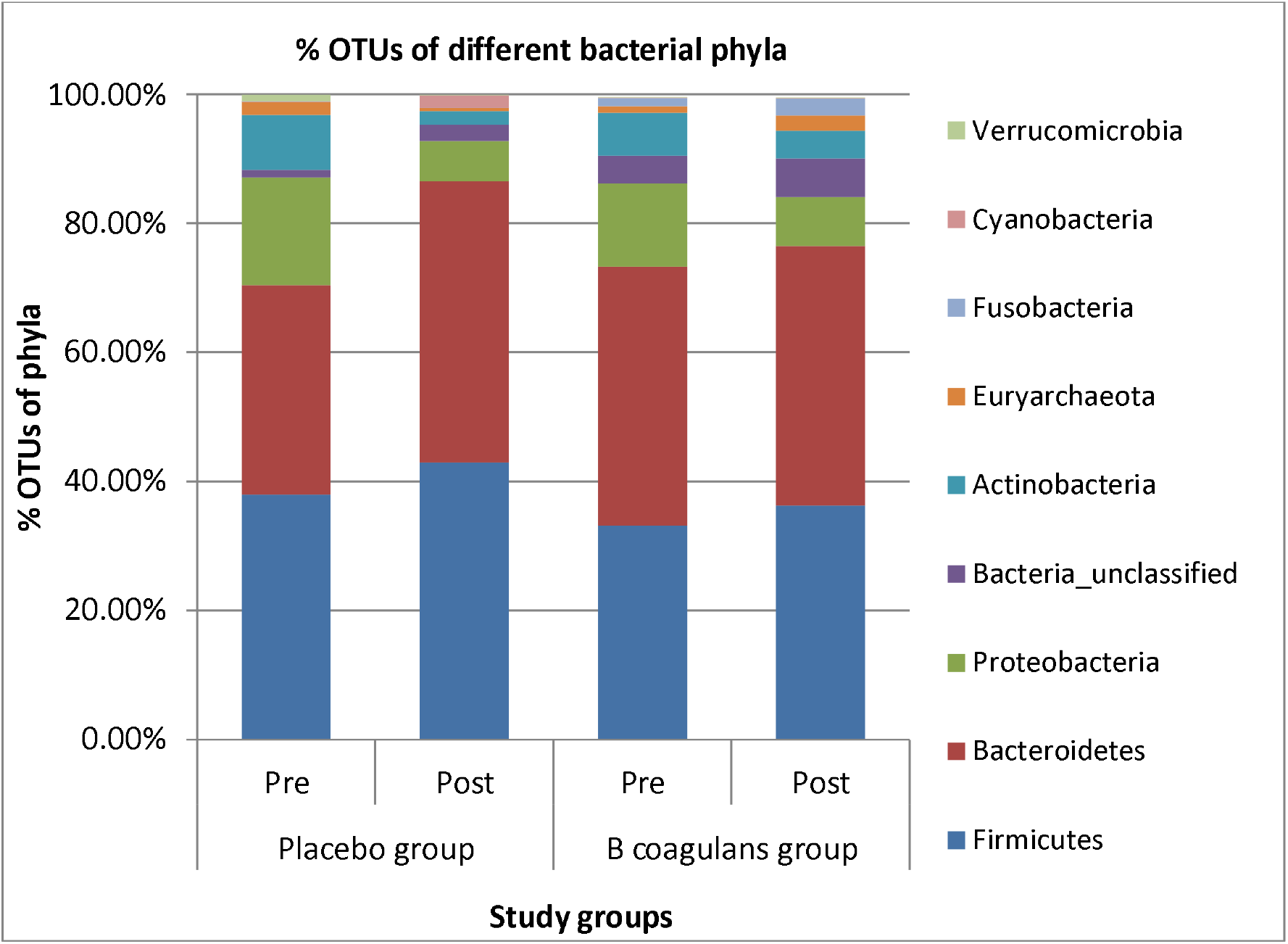
Abundance of major bacterial phyla.

#### Class

The average % OTUs of class Bacteroidia were 40.11% and 40.27% in pre and post intervention sample respectively in treatment group and 32.48% and 43.63% pre and post intervention sample respectively in placebo group. An increase in OTUs of class Bacteroidia was significant in the placebo group. The average % OTUs of class Clostridia were 15.20% and 22.09% in pre and post intervention sample respectively in treatment group and 20.79% and 28.82% in pre and post intervention sample respectively in placebo group. The average % OTUs of class Gammaproteobacteria were 12.87 % and 7.41% in pre and post intervention sample respectively in treatment group and 16.72% and 6.16% respectively in placebo group. The average % OTUs of class Negativicutes were 7.96% and 6.35% in pre and post intervention sample respectively in treatment group and 3.49% and 7.22% respectively in placebo group. A decrease was observed in OTUs of class Negativicutes in post intervention samples of treatment group and an increase in post intervention samples in placebo group. The average % OTUs of class Methanobacteria were 1.02% and 2.36% in pre and post intervention sample respectively in treatment group and 2.03% and 0.55% in placebo group. An increase in OTUs of class Methanobacteria in post intervention samples was observed in treatment group and decrease in OTUs of class Methanobacteria in placebo group. The average % OTUs of class Actinobacteria were 3.49% and 1.81% in pre and post intervention sample respectively in treatment group and 7.76% and 1.64% pre and post intervention sample respectively in placebo group.

**Figure 2:**
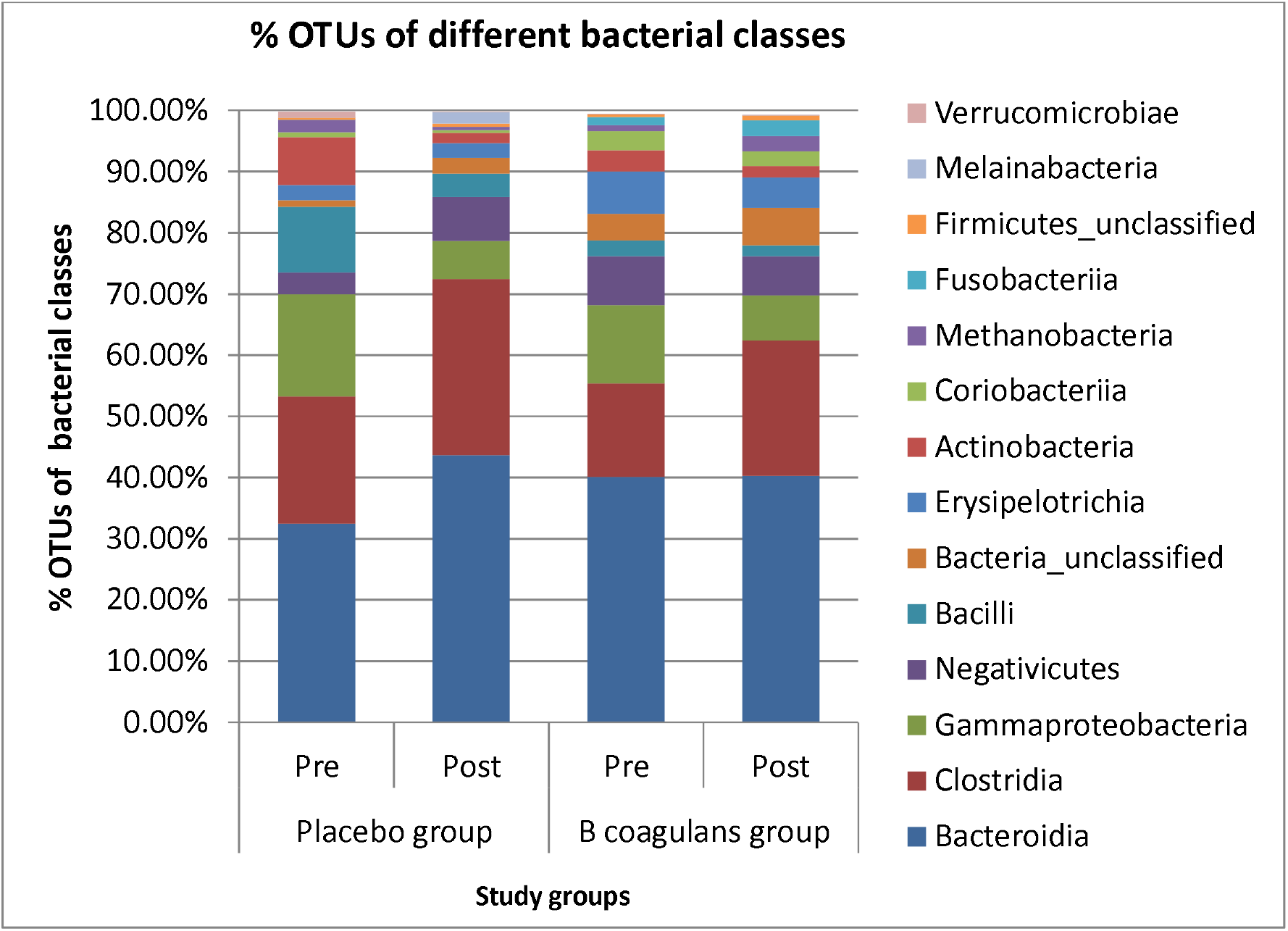
Abundance of major bacterial classes.

#### Order

The average % OTUs of Bacteroidales were 39.94% and 40.03% in pre and post intervention sample respectively in treatment group and 32.37% and 43.46% in placebo group. A significant increase in OTUs of order Bacteroidales in post intervention samples was observed in placebo group. The average % OTUs of order Clostridiales was 15.19% and 22.08% in pre and post intervention sample respectively in treatment group and 20.79% and 28.81% in placebo group. The average % OTUs of order Selenomonadales was 8.02% and 6.30 % in pre and post intervention sample respectively in treatment group and 3.49% and 7.22% in placebo group. A decrease in OTUs of order Selenomonadales in post intervention samples in treatment group and increase in post intervention samples in placebo group. The average % OTUs of order Lactobacillales was 1.67% and 2.49% in pre and post intervention sample respectively in treatment group and 10.66% and 3.82% in placebo group. An increase in OTUs of order Lactobacillales was observed in post intervention samples in treatment group and decrease in post intervention samples in placebo group. The average % OTUs of order Erysipelotrichales was 7.00% and 5.03% in pre and post intervention sample respectively in treatment group and 2.48 % and 2.48% in placebo group. A decrease in OTUs of order Erysipelotrichales in post intervention samples in treatment group and increase in placebo group were observed. The average % OTUs of order Bifidobacteriales were 1.74% and 3.55% in pre and post intervention sample respectively in treatment group and 7.71% and 1.61% respectively in placebo group. An increase in OTUs of order Bifidobacteriales in post intervention samples in treatment group and decrease in placebo group were observed.

**Figure 3:**
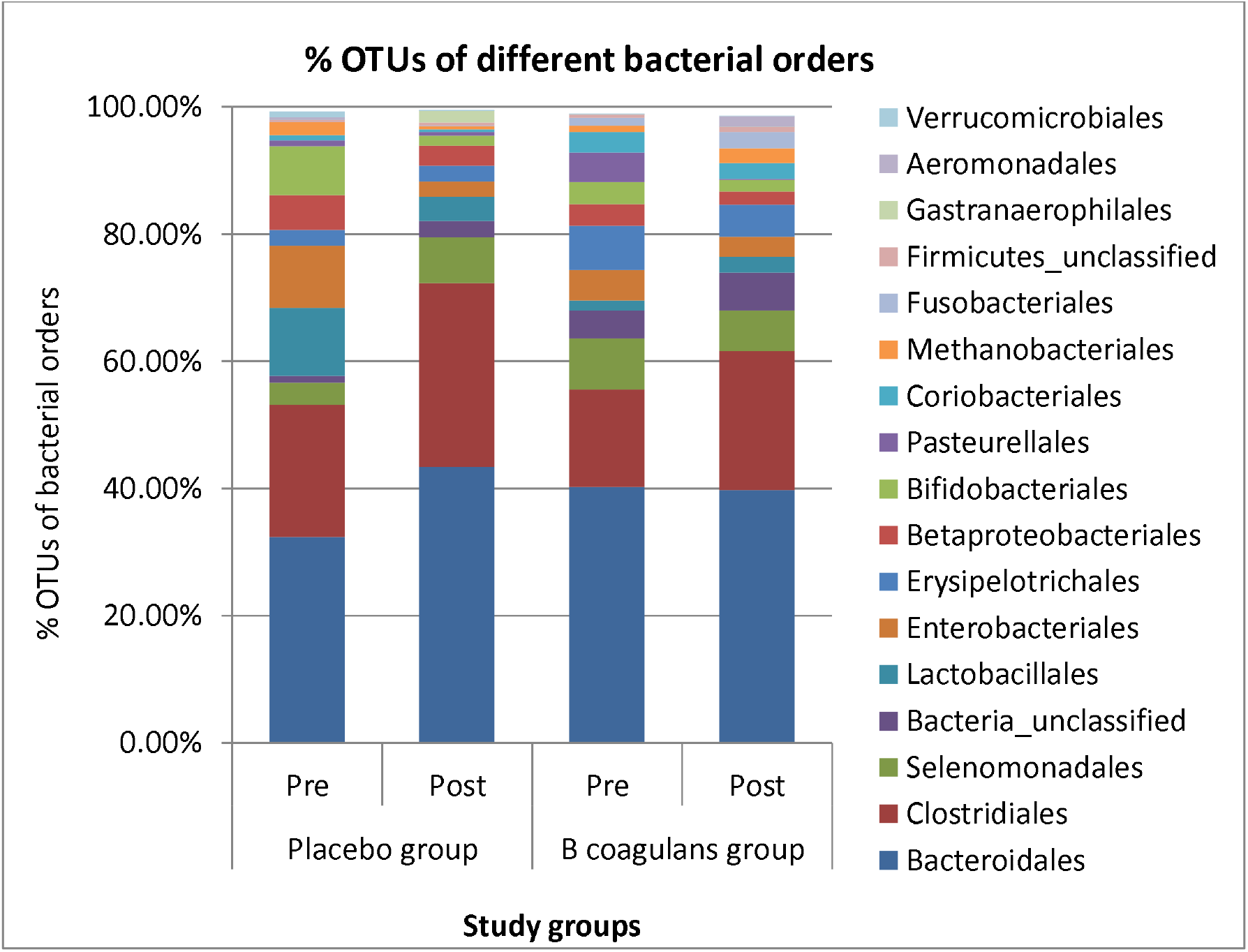
Abundance of major bacterial orders.

#### Family

The average % OTUs of family Prevotellaceae was 32.66% and 33.20% in pre and post intervention sample respectively in treatment group and 8.62% and 31.96% in placebo group. Significant increase was observed in OTUs of family Prevotellaceae in post intervention samples in placebo group. The average % OTUs of family Ruminococcaceae was 8.22% and 11.99% in pre and post intervention sample respectively in treatment group and 12.34% and 12.23% in placebo group. Increase in OTUs of family Ruminococcaceae in post intervention samples in treatment group and decrease in placebo group. The average % OTUs of family Bacteroidaceae was 4.09% and 4.58 % in pre and post intervention sample respectively in treatment group 22.85% and 9.44% in placebo group. Increase in OTUs of family Bacteroidaceae in post intervention samples in treatment group and decrease in placebo group were observed. The average % OTUs of family Lachnospiraceae was 6.14 % and 8.40 % in pre and post intervention sample respectively in treatment group and 6.79 % and 13.48 % respectively in placebo group. The average % OTUs of family Veillonellaceae was 7.47% and 5.72 % in pre and post intervention sample respectively in treatment group and 3.49 % and 7.15% in placebo group. Decrease in OTUs of family Veillonellaceae in post intervention samples in treatment group and increase in placebo group were observed. The average % OTUs of family Enterobacteriaceae was 4.69 % and 3.14 % in pre and post intervention sample respectively in treatment group and 9.79% and 2.41 % respectively in placebo group. A significant decrease was observed in OTUs of family Enterobacteriaceae in post intervention samples in placebo group. The average % OTUs of family Lactobacillaceae was 1.12% and 2.03% in pre and post intervention sample respectively in treatment group and 6.36 % and 3.22 % respectively in placebo group. Increase in OTUs of family Lactobacillaceae in post intervention samples in treatment group and decrease in placebo group were observed. The average % OTUs of family Bifidobacteriaceae was 1.74% and 3.55% in pre and post intervention sample respectively in treatment group and 7.71% and 1.61% respectively in placebo group. Increase in OTUs of family Bifidobacteriaceae in post intervention samples in treatment group and decrease in placebo group were observed.

**Figure 4:**
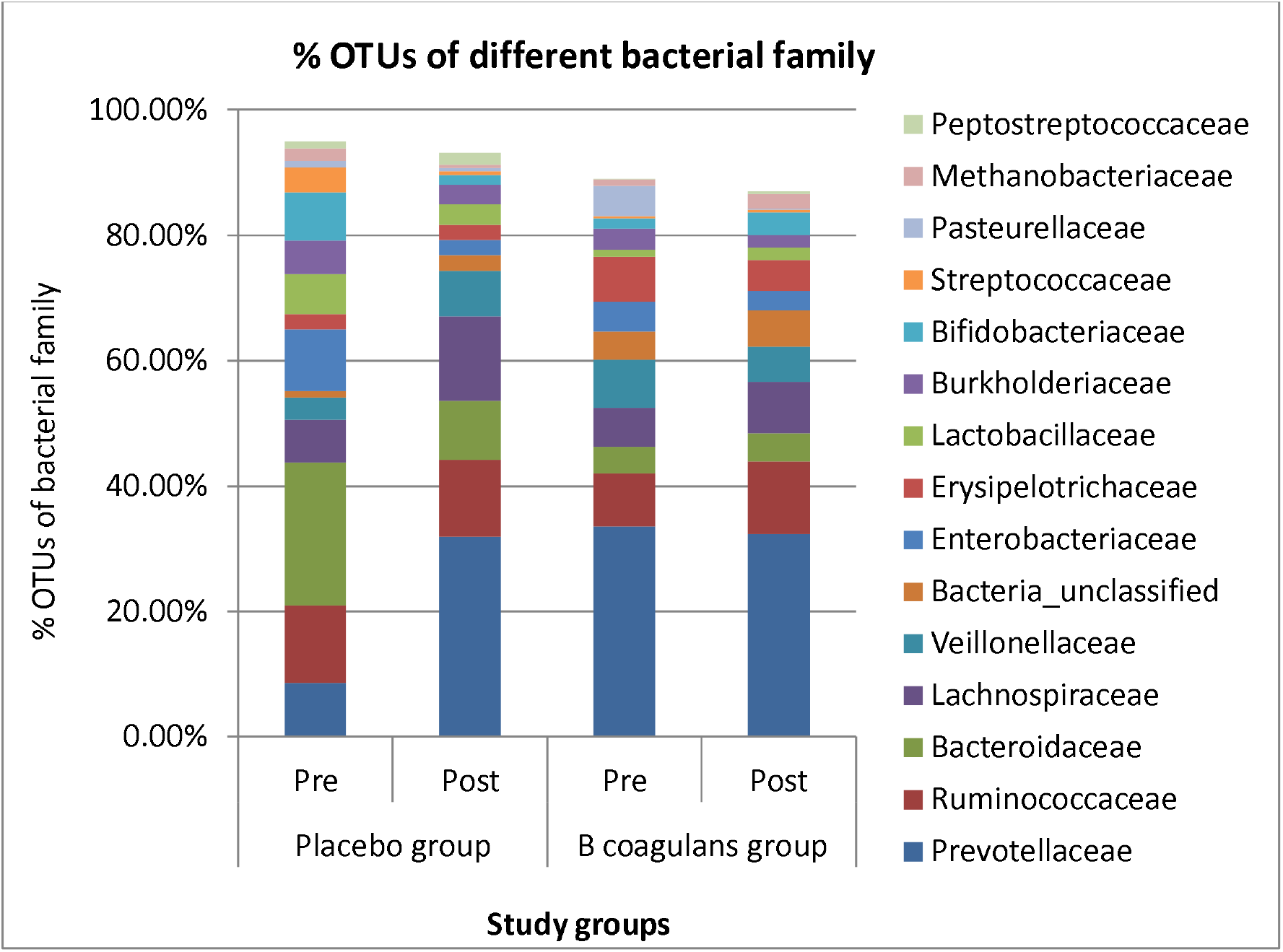
Abundance of major bacterial families.

#### Genera

The average % OTUs of genus *Prevotella* were 26.37% and 22.10% in pre and post intervention sample respectively in treatment group and 10.28% and 25.57% respectively in placebo group. Decrease in OTUs of genus *Prevotella* in post intervention samples in treatment group and an increase in placebo group was observed. The average % OTUs of genus *Bacteroides* were 4.88% and 6.13% in pre and post intervention sample respectively in treatment group and 19.96% and 7.89% respectively in placebo group. An increase in OTUs of genus *Bacteroides* in post intervention samples in treatment group and decrease in placebo group were observed. Average % OTUs of genus *Faecalibacterium* were 6.56% and 6.87% in pre and post intervention sample respectively in treatment group and 9.23% and 8.01% respectively in placebo group. An increase in OTUs of genus *Faecalibacterium* in post intervention samples in treatment group and decrease in placebo group were observed. The average % OTUs of genus *Lactobacillus* were 1.21% and 2.59% in pre and post intervention sample respectively in treatment group and 7.33% and 3.49% respectively in placebo group. An increase in OTUs of genus *Lactobacillus* in post intervention samples in treatment group and decrease in placebo group were observed. The average % OTUs of genus *Bifidobacterium* were 1.67% and 4.44% in pre and post intervention sample respectively in treatment group and 6.03% and 1.42% respectively in placebo group. An increase in OTUs of genus *Bifidobacterium* in post intervention samples in treatment group and decrease in placebo group were observed. The average % OTUs of genus Bacillus were 0.01% and 0.04% in pre and post intervention sample respectively in treatment group and 0.001% and 0.001% respectively in placebo group. OTUs of Genus *Bacillus* in post intervention samples in treatment group were high while OTUs of genus Bacillus in placebo group were very low. The average % OTUs of genus *Escherichia* were 3.61% and 1.24% in pre and post intervention sample respectively in treatment group and 11.06% and 1.81% in placebo group. The average % OTUs of genus *Sutterella* were 3.05% and 2.22% in pre and post intervention sample respectively in treatment group and 3.60% and 1.68 % in placebo group. The average % OTUs of genus *Dialister* were 3.37% and 1.98% in pre and post intervention sample respectively in treatment group and 0.09% and 3.66% respectively in placebo group. A decrease in OTUs of genus *Dialister* was observed in post intervention samples in treatment group and increase in placebo group. The average % OTUs of genus *Megamonas* were 1.39% and 2.17% in pre and post intervention sample respectively in treatment group and 0.73% and 0.001% respectively in placebo group. An increase in OTUs of genus *Megamonas* was observed in post intervention samples in treatment group and decrease in placebo group. The average % OTUs of genus *Roseburia* were 0.82% and 0.45% in pre and post intervention sample respectively in treatment group and 1.18% and 2.46% in placebo group. A decrease in OTUs of genus *Roseburia* was observed in post intervention samples in treatment group and increase in placebo group. The average % OTUs of genus *Megasphaera* were 2.37% and 0.36% in pre and post intervention sample respectively in treatment group and 2.02% and 1.97% in placebo group. A decrease in OTUs of genus *Megasphaera* was observed in post intervention samples in both treatment and placebo group. The average % OTUs of genus *Lachnospira* were 0.42% and 0.34% in pre and post intervention sample respectively in treatment group and 1.49% and 2.83% in placebo group. An increase in OTUs of genus *Lachnospira* was observed in post intervention samples in treatment group and decrease in placebo group. The average % OTUs of genus *Blautia* were 0.25% and 0.40% in pre and post intervention sample respectively in treatment group and 0.12% and 0.25% in placebo group. An increase in OTUs of genus *Blautia* was observed in post intervention samples in both treatment and placebo group. The average % OTUs of genus Alistipes were 0.08% and 0.37% in pre and post intervention sample respectively in treatment group and 0.12% and 0.04% in placebo group. An increase in OTUs of genus Alistipes in post intervention samples in treatment group and decrease in placebo group were observed.

**Figure 5:**
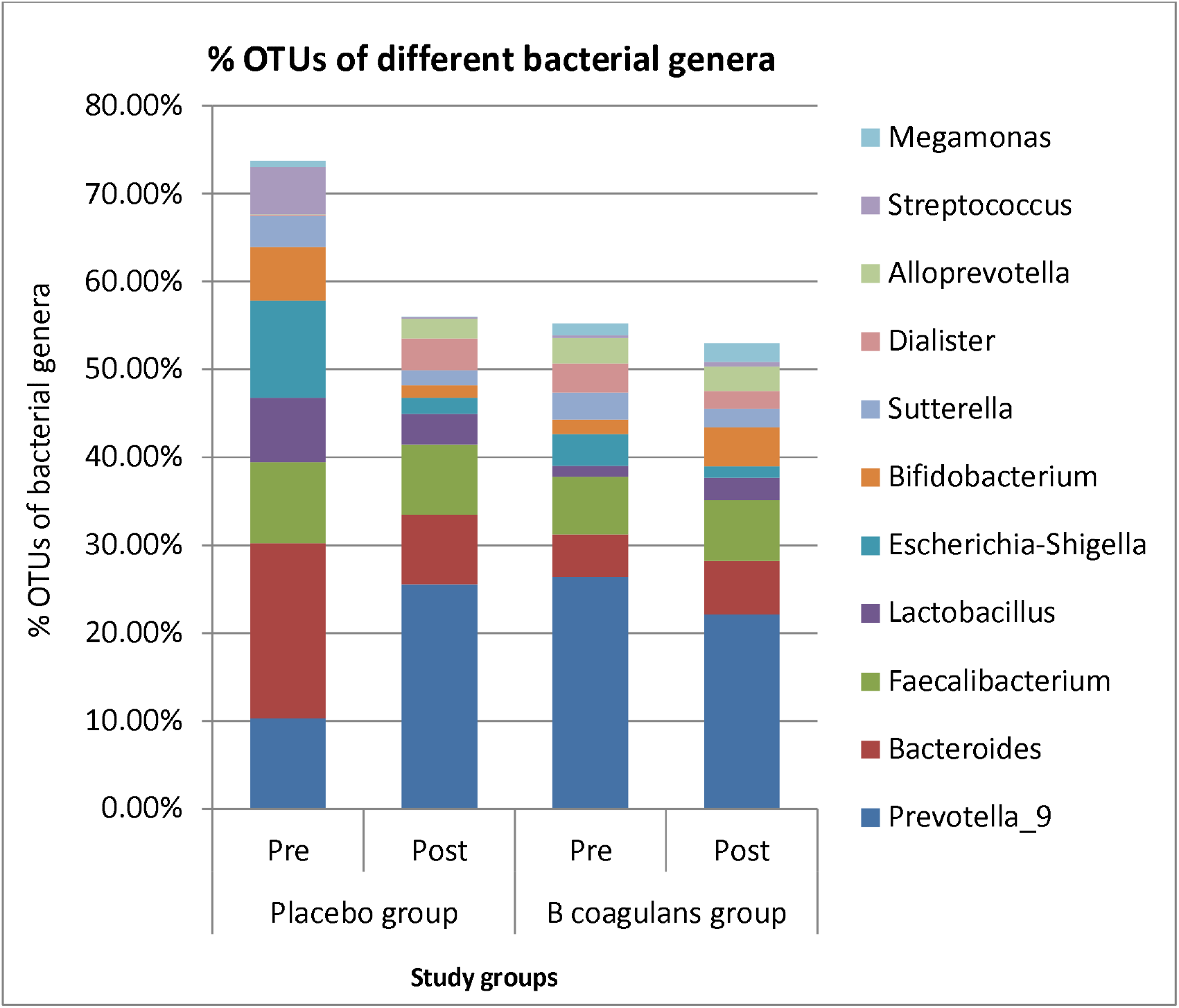
Abundance of major bacterial genera.

### Cytokines levels

Serum concentrations of different cytokines were assessed before and after intervention and results are expressed as Mean ± SD. In the probiotic treated group serum levels of IL10 were 3.23 ± 1.22 pg/ml and 9.25 ± 2.6 pg/ml before and after intervention respectively with the difference being significant (p <0.05). In the placebo group serum IL10 levels were 2.47 ± 1.19 pg/ml and 2.97 ± 1.67 pg/ml before and after intervention respectively, this difference was not significant (p = ns). Results indicated that the secretion of IL-10 in IBD-UC patients was increased in *B coagulans* Unique IS2 group. In the treatment group serum IL6 levels were 34.13 ± 6.8 pg/ml and 17.64 ± 5.2pg/ml before and after intervention respectively with the difference being significant (p <0.05). In the placebo group serum IL6 levels were 35.85 ± 7.6pg/ml and 28.49 ± 6.4pg/ml before and after intervention respectively and this difference was not significant (p = ns). The observed results indicated that the secretion of IL-6 in IBD-UC patients was decreased in *B coagulans* Unique IS2 group. In the probiotic treated group, serum IL17 levels were 42.82 ± 6.9pg/ml and 28.57 ± 5.4pg/ml before and after intervention respectively with the difference being significant (p <0.05). In the placebo group serum IL17 levels were 36.73 ± 13.73 pg/ml and 33.96 ± 14.74 pg/ml before and after intervention respectively with the difference not being significant (p = ns). The observed results indicated that the secretion of serum IL-17 was decreased in IBD-UC patients in the probiotic group. In the probiotic treated group serum IL23 levels were 938.39 ± 56.57 pg/ml and 842.20 ± 69.43 pg/ml before and after intervention respectively with the difference not being significant. In the placebo group serum IL23 levels were 951.59 ± 61.31pg/ml and 932.02 ± 43.30 pg/ml before and after intervention respectively with the difference not being significant (p = ns).

In the treatment group serum IL-1β levels were 358.58 ± 39.29 pg/ml and 267.28 ± 33.88 pg/ml before and after intervention respectively with the difference being significant (p <0.05). In the placebo group serum IL-1β levels were 363.93 ± 35.57 pg/ml and 382.59 ± 37.50 pg/ml before and after intervention respectively with the difference not being significant (p = ns). The observed results indicated that the secretion of serum IL-1β in IBD-UC patients was increased in *B coagulans* Unique IS2 group. In the treatment group serum TNF-α levels were 80.33 ± 13.68 pg/ml and 69.16 ± 14.79 pg/ml before and after intervention respectively with the difference not significant whereas in the placebo group serum TNF-α levels were 76.35 ± 14.72 pg/ml and 79.39 ± 13.8 pg/ml before and after intervention respectively with the difference not being significant.

### Serum serotonin and dopamine levels

In the treatment group serum serotonin level were 121.48 ± 15.52 ng/ml and 111.30 ± 17.74 ng/ml before and after intervention and the difference was not significant. In the placebo group serotonin level were 118.12 ± 19.25 ng/ml and 109.88 ± 11.23 ng/ml before and after intervention and the difference was not significant. In the treatment group serum dopamine level were 8.51 ± 2.52 pg/ml and 11.74 ± 2.25 pg/ml before and after intervention and the difference was not significant. In the placebo group dopamine level were 8.48 ± 2.89 pg/ml and 10.89 ± 2.99 pg/ml, before and after intervention and the difference was not significant. There were changes in serotonin and dopamine levels in the subjects before and after intervention but the difference was not significant.

**Table 1:** Serum Cytokines, serotonin and dopamine levels in pre and post intervention samples in treatment and placebo group.

| Cytokine (pg/ml) | *Bacillus coagulans* Unique IS2 group |  |  | Placebo group |  |  |
| --- | --- | --- | --- | --- | --- | --- |
|  | Pre | Post | P value | Pre | Post | P value |
| IL-10 | $3.23 \pm 1.22$ | $9.25 \pm 2.6$ | <0.05 | $2.47 \pm 1.19$ | $2.97 \pm 1.67$ | NS |
| IL-6 | $34.13 \pm 6.8$ | $17.64 \pm 5.2$ | <0.05 | $35.85 \pm 7.6$ | $28.49 \pm 6.4$ | NS |
| IL-17 | $42.82 \pm 6.9$ | $28.57 \pm 5.4$ | <0.05 | $36.73 \pm 13.7$ | $33.96 \pm 14.74$ | NS |
| IL-23 | $938.39 \pm 56.5$ | $842.20 \pm 69.4$ | NS | $951.59 \pm 61.3$ | $932.02 \pm 43.30$ | NS |
| IL-1β | $358.58 \pm 39.2$ | $267.28 \pm 33.8$ | <0.05 | $363.93 \pm 35.5$ | $382.59 \pm 37.50$ | NS |
| TNF- $\alpha$ | $80.33 \pm 13.6$ | $69.16 \pm 14.7$ | NS | $76.35 \pm 14.7$ | $79.39 \pm 13.8$ | NS |
| Serotonin (ng/ml) | $121.48 \pm 15.5$ | $111.30 \pm 17.7$ | NS | $118.12 \pm 19.2$ | $109.88 \pm 11.23$ | NS |
| Dopamine (pg/ml) | $8.51 \pm 2.52$ | $11.74 \pm 2.2$ | NS | $8.48 \pm 2.89$ | $10.89 \pm 2.99$ | NS |

### Effect on disease symptoms

Symptoms of disease were assessed based on patient complaint and SCCAI score was calculated for IBD-UC patients as per standard protocol. The decrease in the SCCAI score indicates the reduction in the severity of symptoms and increase in the score indicates the augmentation in the severity of the symptom of UC. In this study reduction of 1 value in SCCAI score was considered as decrease in SCCAI score. The SCCAI score was decreased post intervention in 43.75 % of the patients in the probiotic treated group which was significantly high (p <0.05) as compared to placebo where the decrease in SCCAI score was reported in 28.57 % patients.

### Effect on physical, behavioral and psychological parameters

The enrolled subjects were assessed before and after intervention for the different physical symptoms, behavioral and psychological symptoms including stiff or tense muscles, heartburn, headache, shakiness or tremor, sleep problem, difficulty in completing work, procrastination, overwhelming, feeling of depression, trouble relaxing, nervousness, poor concentration, quick temper and restlessness. All the symptoms were evaluated based on scores, the decrease in the score indicates the reduction in the severity of symptoms and increase in the score indicates the augmentation in the severity of the symptom. In this study the complaint of muscles stiffness was reduced post intervention in 41.66 % and 29.16 % subjects in the treatment and placebo group respectively and the difference between the groups was significant (p<0.05). The complaint of heartburn was reduced post intervention in 43.75 % and 31.25 % subjects in the treatment and placebo group respectively and the difference between the groups was significant (p<0.05). The complaints of headache were reduced post intervention in 37.5 % and 33.33 % subjects in the probiotic and placebo group respectively with no significant difference between groups. The complaint of shakiness or tremor was reduced post intervention in 33.33 % and 31.25 % subjects in the treatment and placebo group with no significant difference between groups.

The complaint of sleep problem was reduced post intervention in 41.66 % and 27.08 % subjects in the treatment and placebo group respectively with the difference between groups being significant (p <0.05). The complaint of procrastination was reduced in 31.25 % subjects and 35.4 % subjects in the treatment and placebo group respectively with no significant difference between groups. The complaints of difficulty in completing work or assignments was decreased post intervention in 37.5 % subjects and 27.08 % subjects in the probiotic treated and placebo group respectively with the difference between groups being significant (p <0.05). The complaints of overwhelming was reduced post intervention in 41.66 % and 31.25 % subjects in the treatment and placebo group respectively and the difference between placebo and treatment groups was significant (p <0.05). The complaints of trouble relaxing was reduced post intervention in 37.5 % and 27.08 % subjects in the treatment group and placebo group respectively with the difference between groups being significant (p <0.05). The complaint of nervousness was reduced post intervention in 33.33 % and 29.16 % subjects in the treatment and placebo group respectively with no significant difference between groups. The complaints of depression was reduced post intervention in 33.33 % and 31.25 % subjects in the treatment and placebo group respectively with no significant difference between groups. The complaints of poor concentration was reduced post intervention in 47.91 % and 33.3 % subjects in the treatment group and placebo group respectively with the difference between groups being significant (p<0.05). The complaints of quick temper was reduced post intervention in 45.83 % and 39.58 % subjects in the treatment group and placebo group respectively with no significant difference. The complaints of restlessness was reduced post intervention in 47.91 % and 35.4 % subjects in the treatment group and placebo group respectively with the difference between groups being significant (p<0.05). The observed results exhibited improvement in various physical, behavioral and psychological symptoms of enrolled IBD subjects in the treatment group.

**Table 2:** Post intervention decrease in symptoms in the enrolled subjects for different physical, behavioral and psychological parameters.

| Physical , behavioural and psychological parameters | Post intervention decrease in symptoms (% of total subjects) |  |  |
| --- | --- | --- | --- |
|  | *Bacillus coagulans* Unique IS2 group | Placebo group | P value |
| Muscles stiffness | 41.66 % | 29.16 % | <0.05 |
| Heartburn | 43.75 | 31.25 % | <0.05 |
| Headache | 37.5 % | 33.33 % | NS |
| Shakiness or tremor | 33.33 % | 31.25 % | NS |
| Sleep problem | 41.66 % | 27.08 % | <0.05 |
| Procrastination | 31.25 % | 35.4 % | NS |
| Difficulty in completing work or assignments | 37.5 % | 27.08 % | <0.05 |
| Overwhelming | 41.66 % | 31.25 % | <0.05 |
| Trouble relaxing | 37.5 % | 27.08 % | <0.05 |
| Nervousness | 33.33 % | 29.16 % | NS |
| Depression | 33.33 % | 31.25 % | NS |
| Poor concentration | 47.91 % | 33.3 % | <0.05 |
| Quick temper | 45.83 % | 39.58 % | NS |
| Restlessness | 47.91 % | 35.4 % | <0.05 |

#### Safety evaluations

During and after intervention no adverse events were observed, recorded and reported in the study which further established the safety of *B coagulans* Unique IS2.

## Discussion

Targeted microbiota intervention through probiotics and fecal microbiota transplantation are considered as effective therapeutic methods for IBD^12,33^. Mechanisms of action of probiotics in IBD prevention include increase in beneficial bacteria, inhibition of pathogenic bacteria, immuno-modulation, augmentation of anti-inflammatory responses and enhancement of the intestinal barrier function^14^. *Bacillus* species are high heat resistance, acid tolerance and they can survive considerably better than other probiotics in gastric conditions^34,35^. *B coagulans* Unique IS-2 is a non-toxic commercial probiotic strain with proven safety and efficacy, long shelf life and stability at room temperature^24^. Whole genome sequence analysis of *B. coagulans* Unique IS-2 has corroborated its safety with the absence of any toxin genes^36^. In this study, no severe adverse event was reported, which establish the safety of *B. coagulans* Unique IS-2. Safety and therapeutic efficacy of *B coagulans* Unique IS-2 has been proven in different disease including Irritable Bowel Syndrome in adult^27^ and children^25^, acute-diarrhea^26^, abdominal pain^37^, constipation^27^, oral health^38^, bacterial vaginosis^26^, anti-hypercholesterolemic effect^22^and liver cirrhosis. This strain also showed anti-inflammatory^24^ and anti-proliferative effects^39^.

Post intervention, significant detection of *Bacillus coagulans* in the probiotic treated group demonstrating that this probiotic strain was able to survive in GI tract of IBD patients. Beneficial bacteria provide protection to host against colonization of harmful bacteria and suppress the growth of pathogens by imposing competition for shared niches and nutrients^40^. Results of the present study indicated that *B coagulans* Unique IS-2 was able to modulate the gut microbiota by increasing beneficial bacteria. Another study also reported that consumption of *B. coagulans* was capable of restoring the microbial imbalance^41^ and able to increase populations of *Lactobacillus* and *Bifidobacteria*^42,43^ possibly by consuming free oxygen in the intestine and reduces redox reactions, creating an unfavorable anaerobic and acidic environment to various pathogens^43^.

Results of the metagenomics analysis revealed that phylum Firmicutes, Bacteriodetes, Proteobacteria, Actinobacteria, Euryarchaeota and Verrucomicrobia were abundant in both the study group which are earlier reported dominated in Indian gut^44,45^. An increase in the abundance of phylum Firmicutes and Bacteriodetes was observed in post intervention sample in both groups. Decrease in abundance of phylum Proteobacteria and Actinobacteria was observed in post intervention sample in both groups. Other studies also reported decrease in Bacteroides and Firmicutes and increase in Proteobacteria and Actinobacteria in GI disease^31,46^. It is believed that inflammation is an oxidative state which might promote the outgrowth of aerotolerant taxa such as Proteobacteria and Actinobacteria^47^.

The abundance of *Lactobacillus, Bifidobacterium* and *Bacillus* genera were increased in post intervention samples in treatment group and decrease in placebo group. *Lactobacillus* and *Bifidobacteria* are among the first colonizers of newborns^48^ and are known for their beneficial effects^49^. The observed high abundance of *Lactobacillus, Bifidobacterium* and *Bacillus* genera in treatment group indicated that given probiotic help the gut in restoring these beneficial bacteria. Other studies reported decreased abundance of *Faecalibacterium* in IBD^50,51^. We observed an increase in abundance of genus *Faecalibacterium* in post intervention samples in treatment group and decrease in placebo group which indicates the given probiotic help the gut in restoring them. Bacterial genera *Lactobacillus, Bifidobacterium*, and *Faecalibacterium* have been reported to be protective for mucosal inflammation in the host^11,52^ via several mechanisms, including the up-regulation of the anti-inflammatory cytokine, and down-regulation of inflammatory cytokines^53^. Studies reported decrease in the genera Bacteriodes in IBD^14,54^. We observed an increase in abundance of genus *Bacteroides* in post intervention samples in treatment group and decrease in placebo group which indicates that the given probiotic intervention may help to restore the genus *Bacteroides* in enrolled IBD patients. A decrease in the abundance of bacterial genera *Dialister, Roseburia, Megasphaera* was observed in post intervention samples in treatment group and increase in placebo group. A decrease in the abundance of Blautia species in the IBD patients was reported^55^. We observed an increase in OTUs of genus *Blautia* in post intervention samples in both treatment and placebo group. An increase in abundance of genus Alistipes in post intervention samples in treatment group and decrease in placebo group were observed. Gut microbiota study in the IBD patients reported that some of the Faecalibacterium, *Bacteroides* and Alistipes species have shown significant contribution to metabolic pathway transcription^56^. The abundance of some of bacterial taxon was low but these may play important role in gut function as reported earlier^57^.

The improper host immune response against GI microbiota is considered to be the main reason in causing severe inflammation^40^. Studies have reported the changes in the serum levels of anti-inflammatory cytokine (IL-10) and pro-inflammatory cytokines (IL-6, IL-12, TNF-α, INF-γ) in GI disorders^58^. However, the serum cytokine profiling of IBD patients remains less reported. In the present study significant increase was observed in IL-10 levels in treatment group which indicated that the probiotic strain was able to increase the secretion of IL-10 in IBD patients in the treatment group. Other studies have reported the association of IBD patients with anti-inflammatory cytokines IL-10^59^ and IL-10 secretion increased during disease recovery in IBD patients^60^. It is also reported that inactivation of IL-10 leads to increased release of pro-inflammatory cytokines ^61^.

In the present study we observed the significant decrease in IL6 (p <0.05), IL17 (p <0.05), IL23 (p <0.05), andIL-1β (p <0.05), TNF-α in treatment group which indicated the probiotic intervention was able to modulate the secretion of pro-inflammatory cytokines. Previous studies have reported increased expression of IL-6 may be an intestinal inflammatory mediator of IBD^64^. Studies have reported that the expression of IL-6 was predominantly detected in IBD and an association between serum levels of IL-6 and disease activity^62^. A study reported IL6 in active UC 26 +/- 10 pg/ml and in inactive UC< 10 pg/ml^63^ and this suggested that increased expression of IL-6 may be an intestinal inflammatory mediator of IBD. IL-17 induces the production of many other pro-inflammatory cytokines, including IL-6, TNF-α, and IL-1β, which leads to localizing and amplifying inflammation. IL-17 was reported to be increased in intestinal tissue and serum of IBD patients^64,65^. IL-1β is a pro-inflammatory cytokines play important role in the inflammation in patients with IBD and an elevation in IL-1β levels are associated with increased disease severity^66–68^. Studies reported that the improper level of serotonin and dopamine increases the severity of IBD^69^. Serum serotonin and dopamine were also evaluated in this study but no significant change was observed in both groups.

Results of the study indicated that *B coagulans* Unique IS-2 along with SMT was able to reduce the severity of symptom and improve physical and psychological parameters in IBD patient in the treatment group. These results are similar to another study which reported that a probiotic mixture (VSL#3) reduced the expression of inflammatory cytokines and the severity of disease in UC patients^70^. A meta-analysis also reported that probiotics can benefit IBD treatment during combined use of probiotics and standard therapy^71^. A study with *B. coagulans* Unique IS-2 in children with functional abdominal pain indicated reduction of abdominal pain in the probiotic treated group^37^. Another study reported *B. coagulans* Unique IS-2 was effective in the treatment of IBS with a significant decrease in the intensity of pain in the probiotic treated group^25^. Probiotics can reduce inflammation and disease symptoms by modulation of the mucosal immune system, increased intestinal barrier function, competitive prohibition of pathogens, production of antimicrobial factors^72^ amplification of the intestinal tight junctions to stabilize the permeability, normalize bowel movements and reduce visceral hypersensitivity^73–75^.

## Summary and Conclusion

The results of the study showed that the *B coagulans* Unique IS-2 is able to survive in GI tract of IBD patients. *B. coagulans* Unique IS-2 was able to enhance bacterial genera *Lactobacillus, Bifidobacterium, Faecalibacterium, Bacteroides, Megamonas, Lachnospira, Blautia and* Alistipes in post intervention samples in the treatment group. A decrease in bacterial genera *Sutterella, Dialister, Roseburia and Megasphaera* was observed in post intervention samples in the treatment group. Variable alterations were also observed in the abundance of different bacterial taxon including phylum, class, order, family, and genus in the post intervention sample of the treatment group. *B coagulans* Unique IS-2 was able to modulate the secretion of serum cytokines in IBD patients. The level of IL-10 was increased significantly post intervention in treatment group. The secretion of cytokines, IL-6, IL-1β, TNF-α, IL -17 and IL -23 were variably decreased post intervention in the treatment group. No significant change in serum serotonin and dopamine was observed in both treatment and placebo groups. A reduction in the severity of symptoms of disease and significant improvement in the physical and psychological parameter were observed post intervention in enrolled subjects in the treatment group. Observed results demonstrated that *B coagulans* Unique IS2 showed beneficial effect in IBD-UC patients when administered along with standard medical treatment (SMT). Study was registered with Clinical Trials Registry India (CTRI) - (*CTRI registration No*.*-CTRI/2019/11/022087*).

## Data Availability

The data is included in the manuscript.

## Author Contribution

VDB-Recruited the subjects, carried out experiments, analyzed data and wrote final manuscript, DD - carried out experiments and analyzed the data, PS - carried out experiments and analyzed the data. SK clinical assessment and monitoring of subjects, RSM, JN – drafted study proposal and manuscript, VA-clinical assessment and monitoring of subjects, designed the trial, supervised the study. RC -conceptualizes the study, finalized study proposal, designed the trial and supervised the study. All the authors read and revised the manuscript and approve the final manuscript.

## Acknowledgement

Authors acknowledge the financial support from Unique Biotech Ltd, Hyderabad, India. Help of Mr. Surender Singh (Laboratory Technician) in microbial culture media and reagent preparation is also acknowledged.

